# Long-term Neurocognitive Outcomes after Pediatric Intensive Care Unit Admission: Exploring the Role of Drug Exposure

**DOI:** 10.1101/2022.06.29.22277048

**Authors:** Eleonore S.V. de Sonnaville, Jaap Oosterlaan, Sima A. Ghiassi, Ouke van Leijden, Hanneke van Ewijk, Hennie Knoester, Job B.M. van Woensel, Marsh Königs

## Abstract

**Introduction:** Concerns exist regarding the impact of widely-used clinical drugs on brain development. This study investigates long-term neurocognitive functioning in relation to frequently used drug exposure at the Pediatric Intensive Care Unit (PICU).

**Methods:** This study compared children aged 6-12 years with previous PICU admission (age ≤1 year) for bronchiolitis requiring mechanical ventilation (patient group, n=65) to a demographically comparable control group (n=76) on a broad range of neurocognitive outcomes. The patient group was selected because bronchiolitis seldom manifests neurologically and is therefore not expected to affect neurocognitive functioning in itself. The relation between exposure to sedatives, analgesics and anesthetics and neurocognitive outcomes was assessed by regression analyses.

**Results:** The patient group had lower intelligence than the control group (p<.001, *d*=-0.59) and poorer performance in neurocognitive functions; i.e. speed and attention (p=.009, *d*=-0.41) and verbal memory (p<.001, *d*=-0.60). Exposure to sedatives, analgesics and anesthetics was not related to neurocognitive outcomes.

**Conclusion:** Children with PICU admission for bronchiolitis requiring mechanical ventilation are at risk of long-term neurocognitive impairment. This study found no evidence for a role of exposure to sedatives, analgesics or anesthetics. Findings underline the importance of long-term follow-up after PICU admission, even in absence of disease with neurological manifestation.

**Category of study:** Clinical population study

**Impact:**

- Animal studies have indicated that exposure of the maturing brain to clinical drugs may cause neurodegeneration. Clinical studies show mixed evidence for an association between clinical drugs and neurocognitive outcomes.
- This study provides evidence for long-term neurocognitive impairment among children with a history of PICU admission for bronchiolitis, a condition that seldom manifests neurologically and is therefore not expected to affect neurocognitive functioning in itself.
- We found no evidence for a relation between drug exposure (i.e. sedatives, analgesics and anesthetics) and long-term neurocognitive outcomes, suggesting that the observed neurocognitive impairments are not accounted for by drug exposure.
- Findings underline the importance of structured follow-up after PICU admission, even in absence of disease with neurological manifestation.

## INTRODUCTION

Sedatives, analgesics and anesthetics are routinely used drugs for critically ill children requiring mechanical ventilation at the Pediatric Intensive Care Unit (PICU). Of these drugs, midazolam and morphine are most commonly used and are frequently combined with other sedatives, analgesics or anesthetics.^1-3^ Although long considered to be safe, recent research raises concerns about the potential impact of routinely used drugs on brain development in children.^4^

Animal studies have indicated that exposure to sedatives,^5-7^ analgesics^6^ and anesthetics^5-7^ may cause neurodegeneration, especially in the rapidly developing brain.^5-7^ Several mechanisms are thought to contribute to the potential neurodegenerative impact, such as impaired neurogenesis, reduced synaptogenesis and elevated neuronal apoptosis during early stages of postnatal brain development.^5-11^ Such pathological mechanisms have shown to co-occur with neurocognitive impairments.^5,6,8-10^ Consequently, the US Food and Drug Administration issued a warning for the potential negative impact of repeated and/or longer use of sedatives and anesthetics on brain development in young children.^4^ This raised concerns about the potential impact of routinely used drugs on neurocognitive outcomes of children admitted to the PICU, especially since neurocognitive impairments are known to interfere with development in other major domains of functioning, such as physical and mental health^12,13^, academic achievement^14^, socioeconomic success^15^, and life chances.^13^

A systematic review regarding adult patients presents evidence for a relation between sedative exposure and occurrence of delirium,^16^ while delirium in turn is related to long-term neurocognitive impairment.^17^ Studies directly investigating the relation between exposure to sedatives, analgesics, anesthetics and neurocognitive functioning are scarce. A systematic review^16^ identified two studies reporting a relation between sedative exposure and neurocognitive impairment up to three months after Intensive Care Unit (ICU) stay, while this impairment did not persist at 12-month follow-up.^18,19^ A recent randomized trial in ICU patients revealed no effects of continuous sedation (as compared to no sedation) on neurocognitive functioning at three months after ICU discharge.^20^ Furthermore, some evidence indicates short-term neurocognitive impairment after surgery in adults, although the specific roles of exposure to sedatives, analgesics and anesthetics remain unclear.^21,22^ Taken together, the available literature on adults provides mixed evidence with some indications for short-term effects of sedative exposure on neurocognitive functioning, while no evidence is available regarding longer-term effects of exposure to sedatives, analgesics and anesthetics.

Literature in children is conflicting, with studies showing no relation between exposure to sedatives^23^, analgesics^23^ and anesthetics^24-27^ and neurocognitive outcomes, while other studies do report negative relations with exposure to sedatives^28^, analgesics^29^ and anesthetics^30-33^. Moreover, the available literature is challenged by the unknown contribution of the underlying disease in the observed relations between drug exposure and neurocognitive outcomes.^28,29^ Taken together, it remains unclear to what extent the worrying findings on exposure to sedatives, analgesics and anesthetics in animals - and to some extent in adult patients - generalize to children after PICU admission.

This study investigates neurocognitive outcomes after PICU admission and explores the relation of neurocognitive impairment with exposure to the primary choice of drugs (midazolam and morphine). Secondary analyses also explore relations with exposure to the secondary choice of drugs (lorazepam, fentanyl, esketamine and propofol). We specifically focused on children with bronchiolitis, because this condition seldom manifests neurologically (i.e. 1-2%)^34,35^ and is therefore not expected to affect neurocognitive functioning in itself.

## METHODS

### Participants

This cross-sectional observational study compared a patient group to a control group of peers. The patient group was retrospectively recruited from a cohort admitted between 2007 and 2013 to the PICU of the Amsterdam UMC, The Netherlands. Inclusion criteria for the patient group were: history of PICU admission during infancy or early childhood (age ≤1 year) for respiratory insufficiency due to bronchiolitis requiring invasive mechanical ventilation; age at assessment 6-12 years; and proficient in the Dutch language. Exclusion criteria were: clinical signs of neurological complications (e.g. seizure, encephalitis, meningitis); developmental disorders, physical conditions and/or behavioral deficits interfering with the ability to adequately perform neurocognitive testing; presence of family conflict; and living abroad. Considering the aim of our study, we specifically focused on children with bronchiolitis since we expect minimal involvement of the central nervous system in the pathophysiology. Bronchiolitis is most commonly caused by respiratory syncytial virus (approximately 70% of children hospitalized for bronchiolitis ^36^) that induces cytotoxic injury to lung cells and the subsequent inflammatory response ^37^. Although extrapulmonary manifestations of the infection are well-known,^38^ neurological manifestations are seldom (i.e. 1-2% of cases)^34,35^. Nevertheless, we used clinical signs of neurological manifestations (e.g. seizure, encephalitis, meningitis) as an exclusion criterion in this study. The resulting study sample is relatively well suited for our study aims, since unlike many other diseases treated at the PICU, bronchiolitis in our study sample is not expected to affect neurocognitive functioning in itself.

The control group was recruited using a multichannel approach. Children participating in the patient group were asked to bring a friend or family member. Also, primary schools in the region were contacted for the recruitment of control participants. The inclusion and exclusion criteria for the control group were the same as for the patient group, although children were only included in the control group if they had no history of PICU admission and had not received more than 4 hours sedatives, analgesics and/or anesthetics during their life. We aimed to include at least 64 children in the patient group and 64 children in the control group, in order to achieve sufficient statistical power for the detection of medium-sized effects (Cohen’s *d*=0.5, assuming power=80% and alpha=.05).

### Measures

#### Demographic characteristics

Data on age, sex and socioeconomic status (SES) were collected using a parental questionnaire. SES was estimated by the average level of parental education ranging from 1 (no education) to 8 (postdoctoral education).^39^

#### Clinical characteristics

Administration of sedatives (midazolam and lorazepam), analgesics (morphine and fentanyl) and anesthetics (esketamine and propofol), and clinical characteristics, related to disease severity and with possible impact on neurocognitive functioning, were extracted from the patient files (Table 1). Exposure to each drug was expressed as the total cumulative dose per kilogram bodyweight obtained during PICU admission. Per local clinical protocol at time of PICU admission, the primary choice of drugs during mechanical ventilation consisted of intravenous midazolam and morphine, while the secondary choice of drugs (lorazepam, fentanyl, additional esketamine) were only administered when required. Propofol was only used during (re)intubation and as rescue medication during extreme agitation.

**Table 1.** Demographic and clinical characteristics

| Demographic and clinical characteristics | Patient group<br>(n = 65) | Control group<br>(n = 76) | Mean (SE)<br>difference | p-value | Cohen's *d* |
| --- | --- | --- | --- | --- | --- |
| ***Demographic characteristics*** |  |  |  |  |  |
| Age at time testing (years), mean (SD) | 8.1 (1.2) | 8.2 (1.4) | -0.10 (0.22) | .64 | -0.08 |
| Sex, % boys | 60.0 | 44.7 | 0.63 (0.35) | .08 |  |
| Socioeconomic status, mean (SD) | 5.3 (1.2) | 5.6 (1.0) | 0.01 (0.10) | .92 | 0.01 |
| ***Clinical characteristics*** |  |  |  |  |  |
| Gestational age (weeks), median (IQR) | 38.1 (36.3-39.9) | 39.9 (38.1-40.9) | -0.35 (0.12) * | **.004** | -0.35 |
| Born extremely premature (gestational age <32 weeks), N (%) | 5 (7.7) | 4 (5.3) |  |  |  |
| Age at PICU admission (days), median (IQR) | 43.0 (23.5-79.5) |  |  |  |  |
| Positive respiratory syncytial virus test, N (%) | 56 (86.2) |  |  |  |  |
| PIM 2 score, median (IQR) | 1.4 (1.1-2.1) |  |  |  |  |
| Duration of invasive mechanical ventilation during first PICU stay (hours), mean (SD) | 159.3 (67.9) |  |  |  |  |
| Duration of invasive mechanical ventilation during all PICU admissions (hours), mean (SD) | 169.5 (88.6) |  |  |  |  |
| Length of first PICU stay (days), median (IQR) | 7.4 (5.7-9.0) |  |  |  |  |
| Length of combined PICU stay (days), median (IQR) | 7.6 (5.6-9.4) |  |  |  |  |
| Respiratory syncytial virus positive, N (%) | 56 (86.2) |  |  |  |  |
| Reintubation, N (%) | 4 (6.2) |  |  |  |  |
| Tracheostomy, N (%) | 2 (3.1) |  |  |  |  |
| ECMO, N (%) | 1 (1.5) |  |  |  |  |
| CPR, N (%) | 2 (3.1) |  |  |  |  |
| Readmission at the PICU, N (%) ** | 7 (10.8) |  |  |  |  |
| Withdrawal symptoms, N (%) | 23 (35.4) |  |  |  |  |
| Sepsis (during PICU stay), N (%) | 1 (1.5) |  |  |  |  |
| Septic shock (during PICU stay), N (%) | 0 (0.0) |  |  |  |  |
| Traumatic brain injury after PICU discharge, N (%) | 1 (1.5) | 1 (1.3) |  |  |  |
| ***Sedatives, analgesics and anesthetics during PICU admission*** |  |  |  |  |  |
| Midazolam cumulative mg/kg, median (IQR) | 17.6 (10.7-26.4) |  |  |  |  |
| Morphine cumulative mg/kg, median (IQR) | 1.5 (1.0-2.2) |  |  |  |  |
| Lorazepam, N (%) *** | 17 (26.2) |  |  |  |  |
| Fentanyl, N (%) *** | 20 (30.8) |  |  |  |  |
| Esketamine, N (%) *** | 20 (30.8) |  |  |  |  |
| Propofol, N (%) *** | 16 (24.6) |  |  |  |  |
| Prednisone, N (%) *** | 5 (7.7) |  |  |  |  |
| Dexamethasone, N (%) *** | 44 (67.7) |  |  |  |  |

Note. CPR = cardiopulmonary resuscitation; ECMO = extracorporeal membrane oxygenation; FSIQ = estimated full-scale intelligence quotient; PICU = Pediatric Intensive Care Unit; PIM 2 score = Pediatric Index of Mortality 2 score; SD = standard deviation; SE = standard error.
\* Van der Waerden transformation of gestational age to obtain a normal distribution. \*\* Children were readmitted for viral lower respiratory tract infections and/or subglottic stenosis due to intubation damage. \*\*\* Severely skewed distributions of drug administration, therefore these drugs were dichotomized (i.e. administered yes/no).

#### Intelligence

Intelligence was assessed to capture general neurocognitive functioning and was measured by a short form of the Wechsler Intelligence Scale for Children - Third edition (WISC-III) involving the subtests Vocabulary, Arithmetic, Block Design and Picture Arrangement. Full-scale IQ (FSIQ) estimated with this short form has excellent validity (r=.95) and reliability (r=.90).^40^

#### Neurocognitive domains

In order to assess specific domains of neurocognitive functioning, a standardized and computerized neurocognitive test-battery was used. This test-battery measures a broad range of key neurocognitive domains and contains a composition of child-friendly tests based on well-known neuroscientific paradigms with established validity and reliability, i.e. Attention Network Test,^41^ Multisensory Integration Task,^42^ Tower of London,^43^ Rey Auditory Verbal Learning Test,^44^ Digit Span task,^45^ Klingberg task^46^ and Track & Trace task.^47^ In order to reduce the number of outcome variables, component analysis was used to construct neurocognitive domain scores out of the performance measures resulting from comprehensive neurocognitive assessment (see Supporting Information and eTable 1). The resulting neurocognitive domains and their descriptions are displayed in Table 2.

**Table 2.** Neurocognitive outcomes of children in the patient and control group

| Neurocognitive outcomes | Description | Patient group,<br>Mean (SD)<br>(n = 65) | Control group,<br>Mean (SD)<br>(n = 76) | Mean (SE)<br>difference | p-value | Cohen's *d* |
| --- | --- | --- | --- | --- | --- | --- |
| FSIQ | Intelligence | 95.3 (15.9) | 105.1 (15.1) | -8.46 (1.98) | **<.001** | -0.59 |
| **Neurocognitive domains** |  |  |  |  |  |  |
| Speed and attention | Speed and variability of information processing and attention | -0.19 (0.95) | 0.16 (1.02) | -0.41 (0.15) | **.009** | -0.41 |
| Set shifting | Speed of shifting between response types | -0.03 (1.03) | 0.02 (0.98) | -0.08 (0.16) | .61 | -0.08 |
| Verbal memory | Learning and memory for verbal information | -0.29 (1.13) | 0.24 (0.81) | -0.60 (0.14) | **<.001** | -0.60 |
| Visuomotor integration | Speed and flexibility of visuomotor integration | 0.13 (1.06) | -0.11 (0.93) | 0.25 (0.17) | .14 | 0.25 |
| Verbal working memory | Short-term memory and manipulation of verbal information | -0.15 (1.03) | 0.13 (0.96) | -0.27 (0.16) | .09 | -0.27 |
| Interference control | Speed of suppressing distracting information | 0.12 (1.01) | -0.11 (0.98) | 0.21 (0.16) | .18 | 0.21 |
| Visual processing speed | Speed of visual information processing | -0.03 (1.06) | 0.02 (0.95) | -0.06 (0.16) | .73 | -0.06 |
| Visual working memory | Short-term memory and manipulation of verbal information | -0.16 (1.01) | 0.13 (0.98) | -0.29 (0.17) | .08 | -0.29 |
| Planning time | Speed and capacity of planning ahead | 0.20 (0.99) | -0.17 (0.98) | 0.38 (0.16) | **.02** | 0.38 |
| Multisensory integration | Accuracy for integration of information from different sensory modalities | 0.02 (1.08) | -0.02 (0.93) | 0.04 (0.17) | .82 | 0.04 |

Note. FSIQ = estimated full-scale intelligence quotient; SD = standard deviation; SE = standard error. The directionality of neurocognitive variables was adapted so that for all scores, higher values corresponded to better task performance.

### Procedure

Participating children underwent neurocognitive testing by trained examiners in a quiet room with an approximate duration of three hours, including breaks. Block randomized order of test administration was applied to counterbalance the systematic influence of fatigue on test performance.

### Ethics statement

This study was approved by the medical ethical committee of the Amsterdam UMC (W16_121#16.139) and conducted in accordance with the declaration of Helsinki.^48^ Parents and children aged 12 years provided written informed consent for participation.

### Statistical analysis

Statistical analysis was performed using IBM SPSS Statistics 26.0 and R. Group comparability was tested by comparing the patient and control group on demographic characteristics and gestational age, using mixed modelling to account for the presence of sibling pairs in our sample (*n*=24). Subsequently, groups were compared on the neurocognitive outcomes. For neurocognitive domains with significant group difference, we investigated their more specific nature by following group comparisons on the related original performance measures from neurocognitive tasks (see eTable 1). Neurocognitive outcomes with significant group difference were subjected to subsequent analyses regarding drug exposure.

The primary analysis regarding the relation between drug exposure and neurocognitive outcomes focused on the primary choice of drugs (midazolam and morphine), while the secondary analysis focused on the secondary choice of drugs (lorazepam, fentanyl, esketamine and propofol). We performed univariate regression analyses in the patient group with cumulative dose per kilogram bodyweight as independent variable and neurocognitive outcomes as dependent variables. Skewed distributions of cumulative dose were subjected to logarithmic transformation, while severely skewed distributions were dichotomized (i.e. administered yes/no). Lastly, we explored if exposure to a combination of drugs was related to neurocognitive outcomes. Therefore, we ranked exposure to each drug separately in order of cumulative dose per kilogram bodyweight and calculated the sum of ranks across drugs for each individual child in the patient group. The resulting score reflects the combined exposure to sedatives, analgesics and anesthetics. To correct for multiple testing, correction for false discovery rate (FDR-correction) was applied for all regression analyses other than the primary analysis. All statistical testing was two-sided, α was set at .05 and effect sizes relating to group differences were expressed as Cohen’s *d*.^49^ Cohen’s *d* values of 0.2, 0.5 and 0.8, were used to define thresholds for small, medium and large effect sizes, respectively.^49^

## RESULTS

### Study groups

Children included in the patient group (n=65, Figure 1) did not differ from the total sample of children satisfying the inclusion criteria (n=119) with respect to sex, age at PICU admission, duration of mechanical ventilation and length of PICU stay (eTable 2), indicating no evidence for selection bias in the study sample. Comparisons between the patient and control group (n=76) on baseline characteristics are displayed in Table 1. No differences were found in terms of demographics, indicating no evidence for a confounding role of demographic differences between groups. The patient group had lower gestational age than the control group, of which the role in the results will be explored (see confounding analysis).

**Figure 1.**
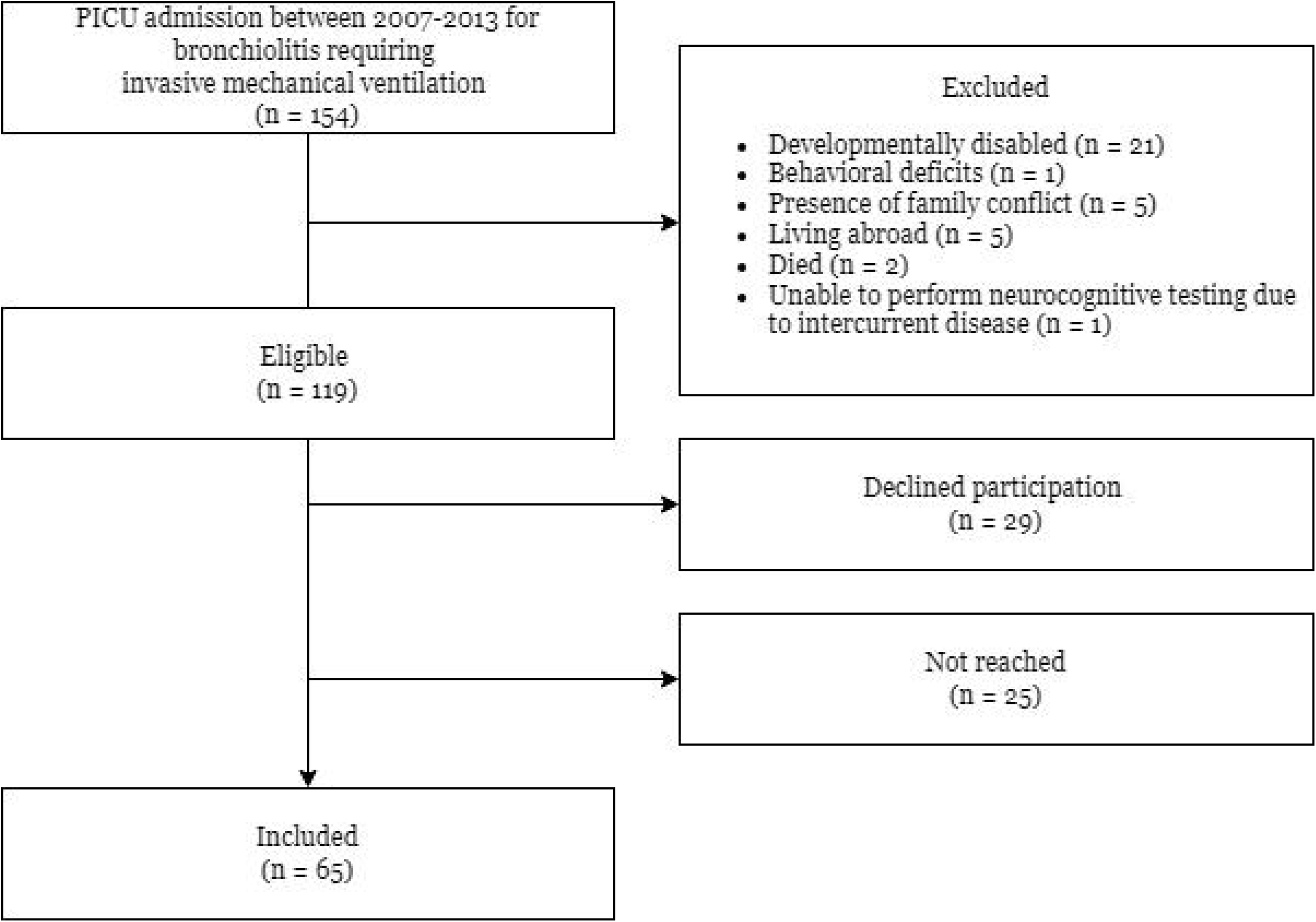
Flowchart of included children in the patient group. Note. Reasons to decline participation were: not interested (n=11), no time (n=10), too high a burden on child (n=6) or language barrier of parents (n=2).

### Group comparison on neurocognitive outcomes

Results regarding neurocognitive outcomes are shown in Table 2. Compared to the control group, the patient group had a significantly lower FSIQ (medium effect), and significant lower performance on the neurocognitive domains for speed and attention (small effect) and verbal memory (medium effect). In contrast, the patient group showed significant better performance than the control group on the neurocognitive domain planning time (small effect). Further analysis of the neurocognitive domain scores at the level of the underlying variables revealed that the observed effect of speed and attention was accounted for by lower processing speed (p=.04, *d*=-0.34), poorer attention consistency (p=.019, *d*=-0.39) and poorer visuomotor accuracy (p=.04, *d*=-0.29) in the patient group. The observed effect for verbal memory was found to be accounted for by poorer verbal memory encoding (p<.001, *d*=-0.61) and poorer verbal memory retrieval (p<.001, *d*=-0.60) in the patient group. While considering impaired verbal memory encoding and retrieval, the patient group had relative better verbal memory consolidation than the control group (p=.03, *d*=0.36). For planning time, the only variable with an effect of group was planning time (p=.006, *d*=0.46), indicating that planning time was shorter in the patient group than in the control group, without a significant difference in planning capacity between the two groups (p=.28, *d*=-0.19).

### Drug exposure and neurocognitive outcomes

We investigated the relation between drug exposure and neurocognitive outcomes, focusing on the neurocognitive outcomes with observed group differences (FSIQ and the neurocognitive domains speed and attention, verbal memory and planning time). Regarding the relation between drug exposure and neurocognitive outcomes (Table 3 and 4), no significant relations were found in the primary analysis (midazolam and morphine), secondary analysis (lorazepam, fentanyl, esketamine and propofol) and tertiary analysis (combined exposure across all drugs).

**Table 3.** Univariate regression analyses testing the relationship of cumulative doses of midazolam and morphine with selected neurocognitive outcomes

| Neurocognitive outcomes | R<sup>2</sup> (%) | Beta (SE) | p-value |
| --- | --- | --- | --- |
| ***Midazolam, cumulative mg/kg*** |  |  |  |
| FSIQ | 0.5 | 1.82 (3.28) | .58 |
| Speed and attention | 0.1 | 0.04 (0.20) | .85 |
| Verbal memory | 0.1 | -0.06 (0.23) | .80 |
| Planning time | 0.4 | 0.10 (0.20) | .62 |
| ***Morphine, cumulative mg/kg\**** |  |  |  |
| FSIQ | 4.2 | 8.90 (5.42) | .11 |
| Speed and attention | 0.6 | -0.21 (0.33) | .53 |
| Verbal memory | 0.2 | 0.15 (0.39) | .71 |
| Planning time | 0.0 | -0.02 (0.34) | .95 |

Note. Beta represents a change of the dependent variable by the independent variable times 10.
\* 1 outlier omitted from analysis.

**Table 4.**
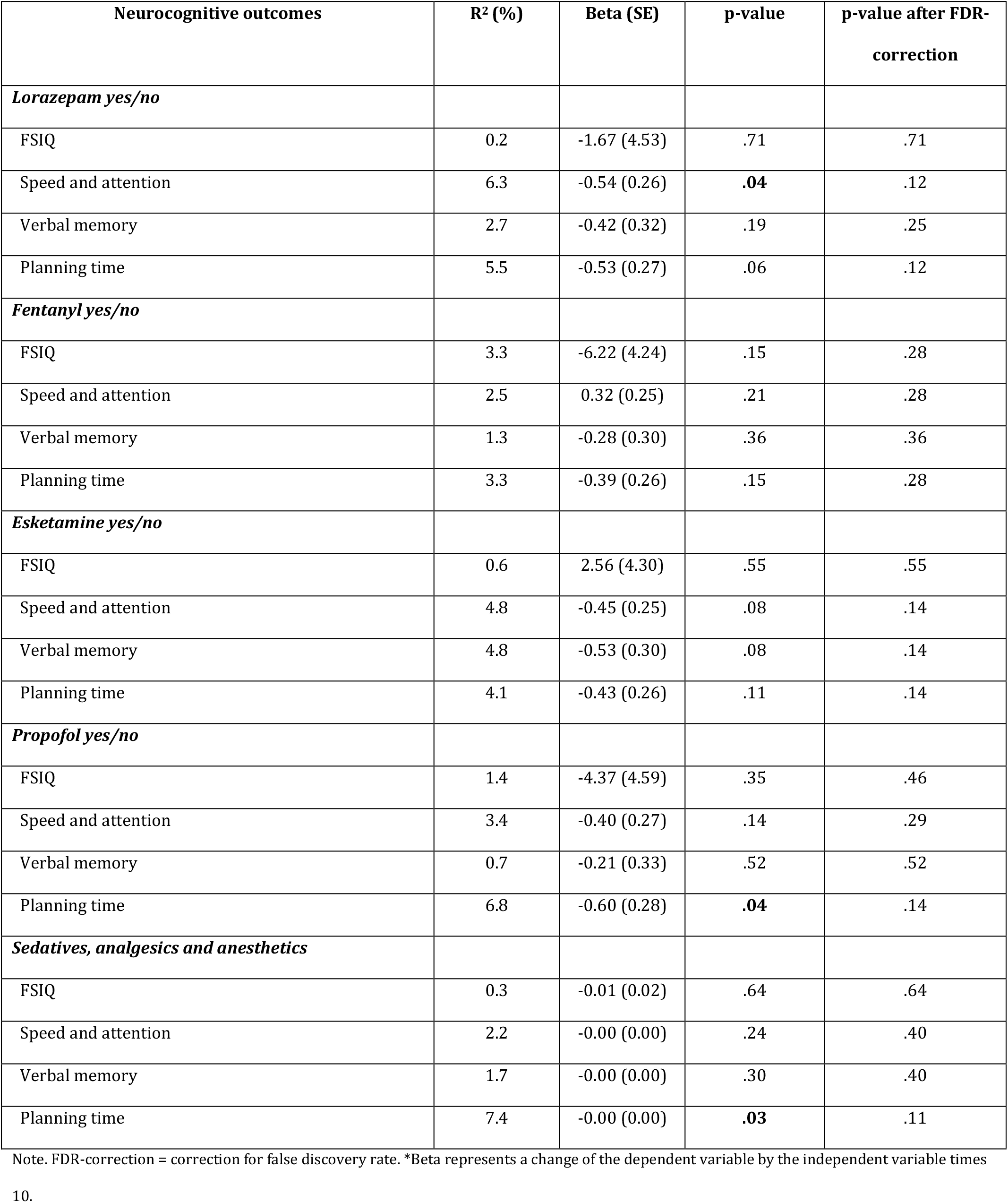
Univariate regression analyses testing the relationship of lorazepam, fentanyl, esketamine, propofol and combination of drugs with the selected neurocognitive outcomes

### Exploratory analysis

As the planned analyses did not reveal relations between drug exposure and neurocognitive outcomes, we performed additional exploratory analyses. In order to exclude the possibility that other aspects of drug exposure than cumulative dose are more important, we additionally performed post-hoc explorations of other aspects of exposure (i.e. duration of administration, mean and highest cumulative day dose), also showing no relations with neurocognitive outcomes (eTable 3). As literature also raised concerns about the potential impact of corticosteroids on neurocognitive functioning in children,^50,51^ we also explored corticosteroid exposure (short course prednisone and dexamethasone to prevent stridor after extubation), and again found no significant relations with neurocognitive outcomes (eTable 3).

### Confounding analysis

As the patient group had significant lower gestational age as compared to the control group, this could theoretically be a confounder in the observed group differences. Therefore, we performed a sensitivity analysis using a subsample of the patient group (n=60) that was comparable to the control group (n=67) in terms of gestational age. The results replicate the reported group differences (ps<.003), indicating that the observed evidence for neurocognitive impairments are not accounted for by premorbid differences in gestational age (see Supporting Information). Various other factors might have accounted for observed group differences. We identified the following relevant factors in the medical history of the patient group: extremely premature birth (gestational age <32 weeks; n=5), CPR (n=2), traumatic brain injury (n=1), septic shock during PICU admission (n=0), ECMO (n=1), and two or more readmissions (n=4). We excluded children with these factors and compared this relatively ‘uncomplicated’ patient subgroup (n=55) to the control subgroup (n=67; eTable 4). Again, we replicated the reported group differences (ps<.024), with the exception of better planning time in the patient group. Taken together, these findings show that the observed evidence for neurocognitive impairments in the patient group is not accounted for by a range of potential confounders.

## DISCUSSION

This study aimed to investigate the relation between sedatives, analgesics and anesthetics and long-term neurocognitive functioning in children with a history of PICU admission. Therefore, we selected a sample of children admitted to the PICU for bronchiolitis, a condition that seldom manifests neurologically (i.e. 1-2%)^34,35^ and is therefore not expected to affect neurocognitive functioning in itself. The results indicate that children with PICU admission for bronchiolitis have affected neurocognitive functioning, reflected by long-term impairment in intelligence and specific aspects of neurocognitive functioning (i.e. information processing, attention, verbal memory and visuomotor integration). Contrary to our hypothesis, exposure to sedatives, analgesics, anesthetics or a combination of these drugs was not related to neurocognitive outcomes. The findings of this study indicate that children admitted to the PICU for bronchiolitis requiring mechanical ventilation are at risk of long-term neurocognitive impairment at primary school age, which is unlikely to be caused by drug exposure during PICU admission.

In 2016, the US Food and Drug Administration warned that repeated or longer use of general sedatives and anesthetics during procedures in children aged less than three years may affect children’s brain development.^4^ As this warning is based on outcomes of animal studies, it remained unclear to what extent these worrying findings could be generalized to children. Studies that reported evidence for potential negative effects of sedatives^28^ and analgesics,^29^ included children in whom the underlying disease is a risk factor for neurocognitive impairment in itself,^52,53^ and drug exposure may have been linked to disease severity in these studies. The findings of this study suggest that exposure to sedatives, analgesics and anesthetics or a combination of these drugs is unlikely to affect long-term neurocognitive outcomes after PICU admission.

The absence of a role for drug exposure in this study raises the question what factors may have contributed to the observed neurocognitive impairments. Other factors may play a role, although we found no evidence for a role of demographic characteristics or medical history (e.g. gestational age, CPR, ECMO). Indeed assuming that bronchiolitis seldom manifests neurologically,^34,35^ the observed neurocognitive impairments may suggest that other pathophysiological mechanisms involving (a combination of) secondary consequences of bronchiolitis and/or PICU treatment may negatively affect neurocognitive outcomes, such as hypoxia, metabolic derangements such as hyponatremia or glucose dysregulation, ischemia, inflammation, hypotension and delirium.^38,54-56^ Likewise, (parental) stress is considered to play an important role after PICU admission^57^ and may be implicated in the mechanisms affecting neurocognitive outcomes.^58,59^ The findings of our study highlight the importance of prospective studies aimed at identifying the combination of factors that may account for neurocognitive impairment in children admitted to the PICU for bronchiolitis, and for PICU admission in general. As neurocognitive impairments are known to interfere with development in crucial outcome domains,^12-15^ our findings also underline the importance of long-term structured follow-up after PICU admission, even in the absence of underlying disease with neurological manifestation, enabling early identification and appropriate management of adverse outcomes.^60^

### Limitations and strengths

This study has several limitations. First, a substantial number of eligible children (45.4%) did not participate in our study, mainly because they were not reached despite our maximal and repeated efforts. Nevertheless, the included children did not differ from the total cohort of eligible children with respect to sex, age at PICU admission, duration of mechanical ventilation and length of PICU stay, indicating no evidence for selection bias in the study sample. Second, the distributions of exposure to lorazepam, fentanyl, esketamine and propofol followed a highly skewed distribution, necessitating dichotomization. This may have reduced the sensitivity of the relevant analyses, although still sufficiently powered to detect medium-sized effects. A strength of this study is the use of a dedicated control group that was comparable to the patient group in terms of age, sex and SES. A comparable control group allows to account for inflation of intelligence over time (known as the Flynn effect)^61,62^ and provides a solution for the inability to correct for SES using standardized norm scores. A second strength is the use of a comprehensive computerized neurocognitive test battery aimed at a broad range of neurocognitive outcomes relevant to daily life functioning. Lastly, we provided a comprehensive analysis of the relation between (combinations of) drug exposure to neurocognitive outcomes.

## CONCLUSIONS

This study provides evidence for long-term neurocognitive impairment among children with a history of PICU admission due to bronchiolitis requiring mechanical ventilation. The results suggest that exposure to sedatives, analgesics and anesthetics is not related to neurocognitive functioning. Future research should aim at identifying factors that are implicated in the neurocognitive impairment of children admitted to the PICU for bronchiolitis. The findings also underline the importance of long-term structured follow-up after PICU admission, even in the absence of underlying disease with neurological manifestation, enabling early identification and appropriate management of adverse outcomes.

## Data Availability

Further group-level data are available on request from the corresponding author. Research data at individual level are not shared due to ethical restrictions under the Dutch law.

## AUTHOR CONTRIBUTIONS

Drs de Sonnaville collected data, carried out the analyses and wrote the manuscript. Prof Oosterlaan, Dr Knoester and Prof van Woensel conceptualized and designed the study and critically reviewed the manuscript. Ms Ghiassi and Mr van Leijden collected data, and reviewed and revised the manuscript. Dr van Ewijk conceptualized and designed the study, collected data, reviewed and revised the manuscript. Dr Königs supervised data collection and analyses, and critically reviewed the manuscript. All authors approved the final manuscript as submitted and agree to be accountable for all aspects of the work.

## ACKNOWLEDGEMENTS

We thank all children and their parents who participated in this study.

## CONSENT STATEMENT

Parents and children aged 12 years provided written informed consent for participation.

## SUPPORT OR GRANT INFORMATION

This study was supported by grants of the Janivo, C.J. Vaillant and Louise Vehmeijer charity foundations. The funder did not participate in the work. This study was further supported by the Emma’s Children Hospital, Amsterdam UMC.

## REFERENCES

1. Minardi C, Sahillioğlu E, Astuto M, Colombo M, Ingelmo PM. Sedation and analgesia in pediatric intensive care. Curr Drug Targets 2012; 13(7): 936–43.

2. Zuppa AF, Adamson PC, Mondick JT, et al. Drug utilization in the pediatric intensive care unit: monitoring prescribing trends and establishing prioritization of pharmacotherapeutic evaluation of critically ill children. J Clin Pharmacol 2005; 45(11): 1305–12.

3. Jenkins IA, Playfor SD, Bevan C, Davies G, Wolf AR. Current United Kingdom sedation practice in pediatric intensive care. Paediatr Anaesth 2007; 17(7): 675–83.

4. FDA Drug Safety Communication: FDA review results in new warnings about using general anesthetics and sedation drugs in young children and pregnant women. 2016.

5. Vutskits L, Xie Z. Lasting impact of general anaesthesia on the brain: mechanisms and relevance. Nat Rev Neurosci 2016; 17(11): 705–17.

6. Istaphanous GK, Ward CG, Loepke AW. The impact of the perioperative period on neurocognitive development, with a focus on pharmacological concerns. Best Pract Res Clin Anaesthesiol 2010; 24(3): 433–49.

7. Zanghi CN, Jevtovic-Todorovic V. A holistic approach to anesthesia-induced neurotoxicity and its implications for future mechanistic studies. Neurotoxicol Teratol 2017; 60: 24–32.

8. Jevtovic-Todorovic V, Hartman RE, Izumi Y, et al. Early exposure to common anesthetic agents causes widespread neurodegeneration in the developing rat brain and persistent learning deficits. J Neurosci 2003; 23(3): 876–82.

9. Lin EP, Soriano SG, Loepke AW. Anesthetic neurotoxicity. Anesthesiol Clin 2014; 32(1): 133–55.

10. Loepke AW, Soriano SG. An assessment of the effects of general anesthetics on developing brain structure and neurocognitive function. Anesth Analg 2008; 106(6): 1681–707.

11. Ikonomidou C, Bosch F, Miksa M, et al. Blockade of NMDA receptors and apoptotic neurodegeneration in the developing brain. Science 1999; 283(5398): 70–4.

12. Koenen KC, Moffitt TE, Roberts AL, et al. Childhood IQ and adult mental disorders: a test of the cognitive reserve hypothesis. Am J Psychiatry 2009; 166(1): 50–7.

13. Gottfredson LS. Why g Matters: The Complexity of Everyday Life. Intelligence; 1997.

14. Petrill SAW B. Intelligence and Achievement: A Behavioral Genetic Perspective. Educational Psychology Review; 2000.

15. Strenze T. Intelligence and socioeconomic success: A meta-analytic review of longitudinal research. Intelligence; 2006. p. 401–26.

16. Kok L, Slooter AJ, Hillegers MH, van Dijk D, Veldhuijzen DS. Benzodiazepine Use and Neuropsychiatric Outcomes in the ICU: A Systematic Review. Crit Care Med 2018; 46(10): 1673–80.

17. Fernandez-Gonzalo S, Turon M, De Haro C, López-Aguilar J, Jodar M, Blanch L. Do sedation and analgesia contribute to long-term cognitive dysfunction in critical care survivors? Med Intensiva 2018; 42(2): 114–28.

18. Jackson JC, Girard TD, Gordon SM, et al. Long-term cognitive and psychological outcomes in the awakening and breathing controlled trial. Am J Respir Crit Care Med 2010; 182(2): 183–91.

19. Pandharipande PP, Girard TD, Jackson JC, et al. Long-term cognitive impairment after critical illness. N Engl J Med 2013; 369(14): 1306–16.

20. Nedergaard HK, Jensen HI, Stylsvig M, et al. Effect of Nonsedation on Cognitive Function in Survivors of Critical Illness. Crit Care Med 2020; 48(12): 1790–8.

21. Ward CG, Eckenhoff RG. Neurocognitive Adverse Effects of Anesthesia in Adults and Children: Gaps in Knowledge. Drug Saf 2016; 39(7): 613–26.

22. Fong HK, Sands LP, Leung JM. The role of postoperative analgesia in delirium and cognitive decline in elderly patients: a systematic review. Anesth Analg 2006; 102(4): 1255–66.

23. Guerra GG, Robertson CM, Alton GY, et al. Neurodevelopmental outcome following exposure to sedative and analgesic drugs for complex cardiac surgery in infancy. Paediatr Anaesth 2011; 21(9): 932–41.

24. Bartels M, Althoff RR, Boomsma DI. Anesthesia and cognitive performance in children: no evidence for a causal relationship. Twin Res Hum Genet 2009; 12(3): 246–53.

25. Sun LS, Li G, Miller TL, et al. Association Between a Single General Anesthesia Exposure Before Age 36 Months and Neurocognitive Outcomes in Later Childhood. Jama 2016; 315(21): 2312–20.

26. Aun CS, McBride C, Lee A, et al. Short-Term Changes in Postoperative Cognitive Function in Children Aged 5 to 12 Years Undergoing General Anesthesia: A Cohort Study. Medicine (Baltimore) 2016; 95(14): e3250.

27. O’Leary JD, Janus M, Duku E, et al. Influence of Surgical Procedures and General Anesthesia on Child Development Before Primary School Entry Among Matched Sibling Pairs. JAMA Pediatr 2019; 173(1): 29–36.

28. Garcia Guerra G, Robertson CM, Alton GY, et al. Neurotoxicity of sedative and analgesia drugs in young infants with congenital heart disease: 4-year follow-up. Paediatr Anaesth 2014; 24(3): 257–65.

29. van Zellem L, Utens EM, de Wildt SN, Vet NJ, Tibboel D, Buysse C. Analgesia-sedation in PICU and neurological outcome: a secondary analysis of long-term neuropsychological follow-up in meningococcal septic shock survivors*. Pediatr Crit Care Med 2014; 15(3): 189–96.

30. Jacola LM, Anghelescu DL, Hall L, et al. Anesthesia Exposure during Therapy Predicts Neurocognitive Outcomes in Survivors of Childhood Medulloblastoma. J Pediatr 2020; 223: 141-7.e4.

31. Backeljauw B, Holland SK, Altaye M, Loepke AW. Cognition and Brain Structure Following Early Childhood Surgery With Anesthesia. Pediatrics 2015; 136(1): e1–12.

32. Zaccariello MJ, Frank RD, Lee M, et al. Patterns of neuropsychological changes after general anaesthesia in young children: secondary analysis of the Mayo Anesthesia Safety in Kids study. Br J Anaesth 2019; 122(5): 671–81.

33. Banerjee P, Rossi MG, Anghelescu DL, et al. Association Between Anesthesia Exposure and Neurocognitive and Neuroimaging Outcomes in Long-term Survivors of Childhood Acute Lymphoblastic Leukemia. JAMA Oncol 2019; 5(10): 1456–63.

34. Pham H, Thompson J, Wurzel D, Duke T. Ten years of severe respiratory syncytial virus infections in a tertiary paediatric intensive care unit. J Paediatr Child Health 2020; 56(1): 61–7.

35. Sweetman LL, Ng YT, Butler IJ, Bodensteiner JB. Neurologic complications associated with respiratory syncytial virus. Pediatr Neurol 2005; 32(5): 307–10.

36. Mansbach JM, Piedra PA, Teach SJ, et al. Prospective multicenter study of viral etiology and hospital length of stay in children with severe bronchiolitis. Arch Pediatr Adolesc Med 2012; 166(8): 700–6.

37. Meissner HC. Viral Bronchiolitis in Children. N Engl J Med 2016; 374(1): 62–72.

38. Eisenhut M. Extrapulmonary manifestations of severe respiratory syncytial virus infection--a systematic review. Crit Care 2006; 10(4): R107.

39. Statistics Netherlands. Education Categorization Standard [Standaard onderwijsinstelling]. Available from: https://www.cbs.nl/nl-nl/onze-diensten/methoden/classificaties/onderwijs-en-beroepen/standaard-onderwijsindeling--soi--/standaard-onderwijsindeling-2006.

40. Sattler JM. Assessment of Children: Cognitive Foundations, 5th Edition. 2008.

41. Fan J, McCandliss BD, Sommer T, Raz A, Posner MI. Testing the efficiency and independence of attentional networks. J Cogn Neurosci 2002; 14(3): 340–7.

42. Königs M, Weeda WD, van Heurn LW, et al. Pediatric traumatic brain injury affects multisensory integration. Neuropsychology 2017; 31(2): 137–48.

43. Shallice T. Specific impairments of planning. Philos Trans R Soc Lond B Biol Sci 1982; 298(1089): 199–209.

44. Saan RJ, & Deelman, B. G.. Nieuwe 15-Woorden Test A en B, 15-WT A en 15-WT B. 1986.

45. Wechsler D. Wechsler Intelligence Scale for Children (3rd ed.) (WISC-III): Manual. San Antonio, TX: The Psychological Corporation.; 1991.

46. Nutley SB, Söderqvist S, Bryde S, Humphreys K, Klingberg T. Measuring working memory capacity with greater precision in the lower capacity ranges. Dev Neuropsychol 2010; 35(1): 81–95.

47. De Kieviet JF, Stoof CJ, Geldof CJ, et al. The crucial role of the predictability of motor response in visuomotor deficits in very preterm children at school age. Dev Med Child Neurol 2013; 55(7): 624–30.

48. World Medical Association Declaration of Helsinki: ethical principles for medical research involving human subjects. Jama 2013; 310(20): 2191–4.

49. Sullivan GM, Feinn R. Using Effect Size-or Why the P Value Is Not Enough. J Grad Med Educ 2012; 4(3): 279–82.

50. Mrakotsky C, Forbes PW, Bernstein JH, et al. Acute cognitive and behavioral effects of systemic corticosteroids in children treated for inflammatory bowel disease. J Int Neuropsychol Soc 2013; 19(1): 96–109.

51. Krull KR, Brinkman TM, Li C, et al. Neurocognitive outcomes decades after treatment for childhood acute lymphoblastic leukemia: a report from the St Jude lifetime cohort study. J Clin Oncol 2013; 31(35): 4407–15.

52. Huisenga D, La Bastide-Van Gemert S, Van Bergen A, Sweeney J, Hadders-Algra M. Developmental outcomes after early surgery for complex congenital heart disease: a systematic review and meta-analysis. Dev Med Child Neurol 2021; 63(1): 29–46.

53. Vermunt LC, Buysse CM, Aarsen FK, et al. Long-term cognitive functioning in children and adolescents who survived septic shock caused by Neisseria meningitidis. Br J Clin Psychol 2009; 48(Pt 2): 195–208.

54. Albin RL, Greenamyre JT. Alternative excitotoxic hypotheses. Neurology 1992; 42(4): 733–8.

55. Johnston MV. Excitotoxicity in perinatal brain injury. Brain Pathol 2005; 15(3): 234–40.

56. Hopkins RO, Jackson JC. Long-term neurocognitive function after critical illness. Chest 2006; 130(3): 869–78.

57. Rees G, Gledhill J, Garralda ME, Nadel S. Psychiatric outcome following paediatric intensive care unit (PICU) admission: a cohort study. Intensive Care Med 2004; 30(8): 1607–14.

58. Malarbi S, Abu-Rayya HM, Muscara F, Stargatt R. Neuropsychological functioning of childhood trauma and post-traumatic stress disorder: A meta-analysis. Neurosci Biobehav Rev 2017; 72: 68–86.

59. Fishbein D, Warner T, Krebs C, Trevarthen N, Flannery B, Hammond J. Differential relationships between personal and community stressors and children’s neurocognitive functioning. Child Maltreat 2009; 14(4): 299–315.

60. Guideline Follow-up of children after admission at the intensive care unit [Richtlijn Follow-up van kinderen na opname op een intensive care]. Available from: https://www.nvk.nl/. 2017.

61. Flynn JR. Are we getting smarter? Rising IQ in the twenty-first century. Cambridge University Press; 2012.

62. te Nijenhuis J, van der Vlier H. Is the Flynn effect on g?: A meta-analysis. Intelligence; 2013. p. 802–7.

