## Supporting Information for "Long-term Neurocognitive Outcomes after Pediatric Intensive Care Unit Admission: Exploring the Role of Drug Exposure"

### Long-term Neurocognitive Outcomes after Pediatric Intensive Care Unit Admission: Exploring the Role of Sedatives, Analgesics and Anesthetics

#### Supporting Information

**Pre-processing**

Missing values (socioeconomic status: 2.1%, neurocognitive data: 1.9%) were imputed using multiple imputation. The neurocognitive test data were subjected to a pre-processing pipeline to construct neurocognitive domain scores out performance measures resulting from comprehensive neurocognitive assessment. First, the directionality of neurocognitive variables was adapted so that for all scores, higher values corresponded to better task performance. Second, to represent all neurocognitive variables on the same scale and reduce the influence of outliers, all variables were subjected to a Van der Waerden transformation.[1] Third, we reduced the number of outcome variables using Principal Component Analysis with oblique rotation.[2,3] The Kaiser criterion was used to determine the number of neurocognitive domains that were selected for further analysis based on the eigenvalue >1.0.[4] Each domain was labeled as a neurocognitive domain based on a selection of variables with the strongest loadings ($-0.5 < r > 0.5$). This procedure resulted in ten neurocognitive domains that explained 78% of the variance contained in the original neurocognitive variables. The neurocognitive domains and the variables corresponding to these domains are displayed in eTable 1.

**eTable 1.** Overview of the neurocognitive domains, variables, definitions and tasks

| **Domains** & Variables | Description | Definition | Task | Description |
|---|---|---|---|---|
| **Speed and Attention** | | | | |
| Processing Speed | The speed of responding to target appearance | Mean reaction time (ms) on trials with neutral targets | Attention Network Test [5] | Target stimuli pointing left or right are presented on a computer screen. Subjects are instructed to respond as quickly as possible to the direction of a target stimulus by pressing the corresponding button. Performance is influenced by the presentation of cues (central, spatial) and manipulation of target flanker congruency (neutral, congruent, incongruent). The measurement of reaction times is corrected for system latency. |
| Processing Stability | The variability of responding to target appearance | Standard deviation of the mean reaction time (ms) on trials with neutral targets | | |
| Attention Consistency | Lapses of attention | The average of the exponential component of the fitted ex-Gaussian curve, reflecting the influence of extremely slow responses (lapses of attention) on information processing | | |
| Visuomotor Accuracy | The precision of proactive visuomotor tracking | The mean distance (in pixels) between the target and the mouse cursor in the structured condition across speed levels | Track & Trace task [6] | See ' Visuomotor Integration' |
| Visuomotor Stability | The variability of proactive visuomotor tracking | The standard deviation of the mean distance (in pixels) between the | | |

| | | | | |
|---|---|---|---|---|
| | | target and the mouse cursor in the structured condition across speed levels | | |
| **Set Shifting** | | | | |
| Speed of set-shifting | The speed of flexibly changing from an automated compatible response to an incompatible response. | The difference in mean reaction time between the set and visual shift trials | Multisensory Integration Task [7] | Measures the ability to flexibly shift between conditions (i.e. set-shifting) and the influence of multisensory integration on set-shifting. In all trials, children were presented with a target in the center of the screen (a penguin) that tilted to the left or to the right, after which a response was required (pressing one of two buttons on a response box). There were three conditions: the *set*, *visual shift* and *audiovisual shift* conditions. In the *set* condition (72% of trials) responses were required to be compatible with the tilt direction of the target. The *visual shift* condition and *audiovisual shift* conditions (14% of trials each) were marked by the presentation of a set-shifting signal at |
| Multisensory integration speed | The speed of integrating information from different sensory modalities. | The difference in mean reaction time between the visual shift trials and audiovisual shift trials | | |

| | | | | |
|---|---|---|---|---|
| | | | | the moment when the target tilted, and required a response that was incompatible with the tilt direction of the target. |
| **Verbal Memory** | | | | |
| Verbal Memory Encoding | The ability to encode verbal information in short-term memory | The sum of correct words recalled over the five direct recall trials | Rey Auditory Verbal Learning Test [8] | A list of 15 words is auditorily presented five times. The subject has to reproduce as many words as possible directly after each presentation (direct recall) and after an interval of 15 minutes (delayed recall). Lastly the subject has to select the presented words among 15 distractors (recognition). |
| Verbal Memory Consolidation | The ability to consolidate verbal information in long-term memory | The difference in the number of correctly recognized words and correctly recalled words in the last direct recall trial | | |
| Verbal Memory Retrieval | The ability to retrieve verbal information from long term memory. | The difference in the number of correctly recognized words and correct words recalled in the delayed recall trial | | |
| **Visuomotor Integration** | | | | |
| Visuomotor Speed | The precision of visuomotor tracking at higher speeds | The difference in mean distance (in pixels) at the highest speed and the lowest speed (across the structured and unstructured condition) | Track & Trace task [6] | A moving target stimulus is presented on the screen of an iPad. Subjects are instructed to keep their index finger on the center of the target in a structured condition (predictable, circular path) and in an unstructured |
| Visuomotor Dynamic Integration | The precision of reactive | The mean distance (in pixels) between the target and the index | | |

| | | | | |
|---|---|---|---|---|
| | visuomotor tracking | finger in the structured condition | | condition (unpredictable, random path) at four linearly increasing target speeds. The speed of the moving stimulus is corrected for the system refreshing rate. |
| **Verbal Working Memory** | | | | |
| Phonological loop | The capacity of encoding visual information in short-term memory. | Performance in the forward condition | Digit Span task [9] | Subjects are required to repeat a sequence of numbers presented auditorily in the order of presentation (forward condition) or reversed order (backward condition). The difficulty increases every other trial, by increasing the length of the sequence of digits. Performance in each condition is defined by the span (the difficulty level of the last correct trial) multiplied by the stability (the total number of correct trials). |
| Verbal Central Executive | The capacity of the central executive to manipulate verbal information in short-term memory. | The difference in performance between the backward and the forward condition | | |
| **Interference Control** | | | | |
| Orienting Attention | The gain in processing speed by spatially orienting attention | The difference in mean reaction time (ms) between trials with spatial and central cues | Attention Network Test [5] | See ‘Speed and Attention’ |

| | | | | |
|---|---|---|---|---|
| Interference Control | The speed of suppressing irrelevant information | The difference in mean reaction time (ms) between trials with incongruent and congruent targets | | |
| **Visual Processing Speed** | | | | |
| Set Speed | The speed of responding to target appearance | Mean reaction time on set trials | Multisensory Integration Task [7] | See 'Set Shifting' |
| Multisensory Integration Speed | The speed of integrating information from different sensory modalities. | The difference in mean reaction time between the visual shift trials and audiovisual shift trials | | |
| **Visual Working Memory** | | | | |
| Visuo-spatial sketchpad | The capacity of encoding visual information in short-term memory | Performance in the forward condition | Klingberg task [10] | A sequence of yellow dots is presented on a four by four digital grid. Subjects are required to repeat the sequence in the order of presentation (forward) or reversed order (backward) by clicking on the relevant locations in the grid. The difficulty increases every other trial, by increasing the length of the sequence or increasing the difficulty of the virtual trajectory of the yellow dots. Performance in each condition is |
| Visual Central Executive | The capacity of the central executive to manipulate visual information in short-term memory. | The difference in performance between the backward and the forward condition | | |

| | | | | |
|---|---|---|---|---|
| | | | | defined by the span (the difficulty level of the last correct trial) multiplied by the stability (the total number of correct trials). |
| **Planning Time** | | | | |
| Alerting Attention | The ability to achieve and maintain an alert state. | The difference in mean reaction time between central cue trials and no cue trials | Attention Network Test [5] | See 'Speed and Attention' |
| Planning Time | The time taken to plan ahead before responding. | Mean of time to first response in trials with correct answers | Tower of London [11] | Colored discs must be moved one by one from an initial state to match a goal state. Instructions are given to plan the whole sequence of moves that must be carried out mentally, before executing the sequence. |
| Planning Capacity | The ability to efficiently plan responses in problem solving | Total items correct multiplied by the maximum correct difficulty degree | | |
| **Multisensory Integration** | | | | |
| Multisensory Integration Accuracy | The accuracy of integrating information from the different sensory modalities. | The difference in mean accuracy between the visual shift trial and audiovisual shift trial | Multisensory Integration Task [7] | See 'Set Shifting' |

Note. Experimental procedures ('Tasks') that have been applied to generate test scores targeting specific neurocognitive functions ('Variables'), which in turn were clustered using component analysis to retrieve overarching scores representing neurocognitive domains ('Domains). ms = milliseconds

**eTable 2.** Comparison of included children with the total sample of eligible children

| Demographic and clinical characteristics | Patient group (n = 65) | Totale sample of eligible children (n = 119) | p-value |
|---|---|---|---|
| Sex, % boys | 60.0 | 59.7 | .08 |
| Age at PICU admission (days), median (IQR) | 43.0 (23.5-79.5) | 45.0 (27.0-82.0) | .56 |
| Mechanical ventilation (days), mean (SD) | 6.6 (2.8) | 6.3 (2.7) | .27 |
| PICU stay (days), median (IQR) | 7.4 (5.7-9.0) | 6.88 (5.0-8.7) | .23 |

Note. PICU = Pediatric Intensive Care Unit

**eTable 3.** Exploratory analysis

| Neurocognitive outcomes | $R^2$ (%) | Beta (SE) | p-value | p-value after FDR-correction |
|---|---|---|---|---|
| ***Invasive mechanical ventilation duration*** | | | | |
| FSIQ | 0.2 | 0.01 (0.02) | .69 | .69 |
| Speed and attention | 0.3 | -0.00 (0.00) | .69 | .69 |
| Verbal memory | 2.3 | -0.00 (0.00) | .23 | .69 |
| Planning time | 0.4 | -0.00 (0.00) | .60 | .69 |
| ***Midazolam mean cumulative daydose*** | | | | |
| FSIQ | 0.5 | 2.10 (3.90)* | .59 | .79 |
| Speed and attention | 0.3 | 0.09 (0.23)* | .69 | .79 |
| Verbal memory | 0.1 | 0.07 (0.28)* | .79 | .79 |
| Planning time | 0.7 | 0.17 (0.24)* | .50 | .79 |
| ***Midazolam highest cumulative daydose*** | | | | |
| FSIQ | 1.6 | 3.90 (4.18)* | .36 | .54 |
| Speed and attention | 8.5 | 0.48 (0.22)* | **.03** | .13 |
| Verbal memory | 1.3 | 0.23 (0.28)* | .41 | .54 |
| Planning time | 0.1 | 0.07 (0.27)* | .78 | .78 |
| ***Morphine mean cumulative daydose*** | | | | |
| FSIQ | 4.4 | 10.62 (6.29)* | .10 | .34 |
| Speed and attention | 0.3 | -0.17 (0.38)* | .65 | .87 |
| Verbal memory | 3.0 | 0.62 (0.44)* | .17 | .34 |
| Planning time | 0.0 | 0.03 (0.40)* | .93 | .93 |
| ***Morphine highest cumulative daydose*** | | | | |
| FSIQ | 0.0 | 0.63 (7.21)* | .93 | .93 |
| Speed and attention | 1.0 | -0.27 (0.39)* | .49 | .65 |
| Verbal memory | 1.5 | -0.42 (0.47)* | .38 | .65 |
| Planning time | 1.1 | -0.33 (0.45)* | .46 | .65 |
| ***Prednisone yes/no*** | | | | |
| FSIQ | 0.0 | 0.00 (7.47) | .99 | .99 |
| Speed and attention | 0.7 | 0.30 (0.44) | .50 | .97 |
| Verbal memory | 1.6 | -0.52 (0.53) | .32 | .97 |
| Planning time | 0.2 | -0.16 (0.47) | .73 | .97 |

| | | | | |
|---|---|---|---|---|
| ***Dexamethasone yes/no*** | | | | |
| FSIQ | 2.0 | -4.73 (4.22) | .27 | .45 |
| Speed and attention | 1.6 | -0.25 (0.25) | .32 | .45 |
| Verbal memory | 1.5 | 0.29 (0.30) | .33 | .45 |
| Planning time | 0.1 | 0.08 (0.27) | .77 | .77 |
| ***Sedatives, analgesics, anesthetics and corticosteroids*** | | | | |
| FSIQ | 0.4 | -0.01 (0.02) | .60 | .60 |
| Speed and attention | 1.6 | -0.00 (0.00) | .31 | .51 |
| Verbal memory | 1.2 | -0.00 (0.00) | .38 | .51 |
| Planning time | 5.2 | -0.00 (0.00) | .07 | .27 |

Note. FDR-correction = correction for false discovery rate. *Beta represents a change of the dependent variable by the independent variable times 10.

#### Confounding analysis

As the patient group had significant lower gestational age as compared to the control group, this could theoretically be a confounder in the observed differences between the patient and control group. Therefore, we assessed whether the neurocognitive variables that were significantly different between the patient and control group, were also related to gestational age. This was the case for FSIQ (p=.030) and for verbal memory (p=.002), but not for speed and attention (p=.071) nor for planning time (p=.124). In order to create a patient and control group comparable on gestational age, we excluded 5 children in the patient group with gestational age <32 weeks en 9 children in the control group with gestational age >41.5 weeks (median (IQR) respectively 38.36 (36.89-40.11) weeks and 39.57 (38.00-40.43) weeks, p=.259). Subsequently, we repeated the group comparisons for FSIQ and verbal memory, which replicated the previously reported significant group differences (mean (SE) difference respectively -8.46 (2.24), p<.001 and -0.47 (0.15), p=.003). These findings indicate that the observed evidence for neurocognitive impairments are not accounted for by premorbid differences in gestational age.

**eTable 4.** Confounding analysis

| Demographic and clinical characteristics and neurocognitive outcomes | mean (SE) difference between patient and control group | p-value |
|---|---|---|
| ***Demographic and clinical characteristics*** | | |
| Age at time testing (years) | -0.20 (0.24) | .41 |
| Sex (% boys) | 0.57 (0.37) | .13 |
| Socioeconomic status | 0.01 (0.12) | .96 |
| Gestational age (weeks)* | -0.15 (0.12) | .23 |
| ***Neurocognitive outcomes*** | | |
| FSIQ | -7.71 (2.25) | **.001** |
| Speed and attention | -0.40 (0.17) | **.02** |
| Set shifting | -0.18 (0.18) | .32 |
| Verbal memory | -0.44 (0.15) | **.005** |
| Visuomotor integration | 0.25 (0.18) | .16 |
| Verbal working memory | -0.27 (0.17) | .12 |
| Interference control | 0.33 (0.16) | .05 |
| Visual processing speed | -0.12 (0.17) | .49 |
| Visual working memory | -0.30 (0.18) | .11 |
| Planning time | 0.32 (0.18) | .08 |
| Multisensory Integration | 0.04 (0.18) | .83 |

Note. Patient group n = 55, control group n = 67: Excluded in patient group: gestational age <32 weeks, bronchopulmonary dysplasia, cardiopulmonary resuscitation, traumatic brain injury, septic shock, delirium, Pediatric Index of Mortality 2 score >10, extra-corporeal membrane oxygenation, more than two PICU admissions. Excluded in control group: gestational age >41.5 weeks to have a comparable gestational age between patient and control group. * Van der Waerden transformation of gestational age to obtain a normal distribution.

#### References


1. Van Der Waerden B. Mathematical statistics springer–verlag. ew York; 1969.
2. Holland SM. Principal components analysis (pca). 2008.
3. Rummel RJ. Applied factor analysis. Northwestern University Press; 1988.
4. Kaiser HF. The application of electronic computers to factor analysis. Educational and psychological measurement; 1960. p. 141–51.
5. Fan J, McCandliss BD, Sommer T, Raz A, Posner MI. Testing the efficiency and independence of attentional networks. *J Cogn Neurosci* 2002; **14**(3): 340-7.
6. De Kieviet JF, Stoof CJ, Geldof CJ, et al. The crucial role of the predictability of motor response in visuomotor deficits in very preterm children at school age. *Dev Med Child Neurol* 2013; **55**(7): 624-30.
7. Königs M, Weeda WD, van Heurn LW, et al. Pediatric traumatic brain injury affects multisensory integration. *Neuropsychology* 2017; **31**(2): 137-48.
8. Saan RJ, & Deelman, B. G. . Nieuwe 15-Woorden Test A en B, 15-WT A en 15-WT B. 1986.
9. Wechsler D. Wechsler Intelligence Scale for Children (3rd ed.) (WISC-III): Manual. San Antonio, TX: The Psychological Corporation.; 1991.
10. Nutley SB, Söderqvist S, Bryde S, Humphreys K, Klingberg T. Measuring working memory capacity with greater precision in the lower capacity ranges. *Dev Neuropsychol* 2010; **35**(1): 81-95.
11. Shallice T. Specific impairments of planning. *Philos Trans R Soc Lond B Biol Sci* 1982; **298**(1089): 199-209.
